# OX2: Understanding the pattern of symptoms for pancreatic neuroendocrine neoplasms (PNENs) to improve the early diagnosis of pancreatic cancer: analysis of the QResearch database

**DOI:** 10.1101/2021.02.03.21250038

**Authors:** Investigators, Weiqi Liao, Ashley Clift, Martina Patone, Carol Coupland, Julia Hippisley-Cox

## Abstract

This article is the study protocol and statistical analysis plan for the project. This study is set up to identify and quantify “red flag” symptoms associated with the diagnosis of Pancreatic Neuroendocrine Neoplasm (PNEN) and compare these with the symptoms associated with Pancreatic Ductal Adenocarcinoma (PDAC). The results of this study will inform the evidence base for the refinement of the QCancer (Pancreas) tools, which have been integrated into the UK NHS primary care computer systems, to improve the early recognition of PNEN.

## 2 Research background

Pancreatic cancer is the 10^th^ most common cancer in the UK^1^. Diagnoses are often made late when the cancer is advanced^2^. Less than 20% of patients are suitable for surgery and about 75% of patients are likely to die within a year of diagnosis^1^, which is the worst survival rate for any cancer^2^. However, better survival is more likely if patients present at an early stage^1^. There are only a few established risk factors for pancreatic cancer such as age^2^, smoking^2,3^, genetic factors^2^, chronic pancreatitis^1^ and alcohol^1,4^. Diabetes may be a risk factor for pancreatic cancer or an early manifestation of a growing tumour^5,6^. It is unlikely that there will be a national screening programme for pancreatic cancer since there are few established risk factors and currently no reliable screening test. Hence, it is likely that most pancreatic cancers will be diagnosed in symptomatic patients presenting to primary care. The challenge then becomes enabling earlier diagnosis to help improve treatment options (e.g. possibility of surgery) and prognosis. Earlier diagnosis could be helped by increased public awareness of symptoms which might indicate pancreatic cancer such as weight loss, loss of appetite, and abdominal pain^3,6,7^. It could also be improved by more prompt investigation of symptomatic patients presenting to their GP^6^. In the UK, GPs will soon have better direct access to diagnostic investigations such as ultrasound, CT scan, and MRI. GPs need better assessment tools to quantify a patients risk of different types of cancer so that the right patients are sent for the right investigations, and making efficient use of scarce resources^6^.

There are two types of pancreatic cancer tumour

1. Exocrine tumours start in the exocrine cells of the pancreas, where enzymes which help digest food are made. About 95 out of 100 (95%) pancreatic cancers are exocrine tumours.
2. Neuroendocrine tumours that start in the pancreatic endocrine cells, which produce hormones.

Pancreatic neuroendocrine neoplasms (PNENs) start in the cells in the pancreas that produce hormones, called endocrine cells or Islet of Langerhans cells. Neuroendocrine tumours (NETs) start in the neuroendocrine system throughout the body. The neuroendocrine system produces hormones. Neuroendocrine tumours can be non-cancerous (benign), or cancerous (malignant). The pancreatic endocrine cells produce hormones, including insulin and glucagon. About one or two in a hundred pancreatic cancers (1-2%) are PNENs with the remainder being pancreatic adenocarcinoma (PDAC). 10-30% of pancreatic neuroendocrine tumours produce specific hormones, which can cause specific symptoms (functioning PNENs). They can cause symptoms such as pain, weight loss, jaundice, or diarrhoea, which are similar to the symptoms of pancreatic ductal adenocarcinoma (PDAC). Non-functioning pancreatic neuroendocrine tumours may be harder to detect and diagnose than functioning neuroendocrine tumours because they don’t cause specific symptoms, but there are no large-scale studies documenting this in detail, which is the research gap this study aims to address.

## 3 Aim and objectives of this study

This study aims to identify and quantify ‘red flag’ symptoms associated with a diagnosis of PNEN, and compare these with the symptoms associated with PDAC tumours. This study will inform the evidence base for the refinement of the QCancer tools, which have been integrated into the GP systems to improve the early recognition of PNENs.

The specific research objectives of this study are:

1. To identify and describe the characteristics of patients with a diagnosis of PNEN using the QResearch database linked to cancer registry, mortality and hospital data;
2. To identify which of the known red flag symptoms are significantly and independently associated with a diagnosis of PNENs;
3. To provide information to feed into the health economics model where appropriate.

## 4 Methods

### 4.1 Study design

Case control and cohort studies in primary care.

### 4.2 Study period

01 Jan 2000 to 31 December 2019

### 4.3 Setting and data source

QResearch is a large validated research database including the records of approximately 35 million patients registered with approximately 1500 GP practices using the EMIS Health Clinical System including data since 1989 offering longitudinal data for over 20 years. It is the largest database of its kind and more than twice the size of CPRD. The QResearch database includes event level detailed information on patient demographics (year of birth, sex, ethnicity, deprivation), medication, clinical diagnoses, referrals, clinical values, laboratory investigations. All 1500 practices have been linked at individual patient level to cancer registrations data (from 1990 onwards), mortality records (from 1997 onwards) and to hospital admissions data (from 1998 onwards)^8^. The mortality records include cause of death (up to 15 causes). The cancer registration data includes information on the date of diagnosis, type and location of the tumour, morphology, grade and stage, treatment (surgery, chemotherapy, radiotherapy). QResearch is representative of the UK population. The database has been used extensively for research including cancer epidemiology, health services research, the development of risk prediction models. It has an excellent track record of high impact research papers published in leading journals including studies using the linked datasets^9-13^. It was used to develop the QCancer tools to improve the early diagnosis of cancer which are now implemented into the majority of GP practice computer systems^9,10,12^.

### 4.4 Study population

An open cohort of adult patients aged ≥25 years registered during the study period 1^st^ January 2000 to 31^st^ December 2019 will be identified for this study. Patients with an existing diagnosis of any type of pancreatic cancer at the start of the study will be excluded. The entry date to the study cohort will be the latest of 25^th^ birthday, date of registration with the practice plus one year, date on which the practice computer system was installed plus one year, or the beginning of the study period (1^st^ January 2000). The right censor date will be the earliest of the following: date of diagnosis of PNEN, date of death, date of leaving the practice, date of the latest download of data or the study end date (31^st^ December 2019). Person years will be calculated between the study entry date and the right censor date.

### 4.5 Identification of cases

Cases will be patients in the study cohort with a new diagnosis of pancreatic neuroendocrine tumour plus another set of cases with PDAC cancer, on either GP, hospital records, cancer registration data or deaths between 2000 and 2019. A feasibility analysis identified about 620 patients with PNEN on QResearch since 1998. Where possible, cases will be classified into main types of PNENs, i.e. gastrinomas, insulinomas, glucagonomas, somatostainomas, and VIPomas.

### 4.6 Identification of controls

Controls will be patients at the same age and sex as the case but without a diagnosis of PDAC or PNEN. Up to 10 controls who are alive and registered at the time of the diagnosis of the case (index date) will be matched with each case. Controls will be matched with cases by practice, age, sex, and calendar time using incidence density sampling^14^. Each control will be allocated an index date which will be the date of cancer diagnosis for their matched case.

### 4.7 Shortlisting candidate red flag symptoms

A list of ‘red flag’ symptoms will be identified for this study. Starting with those already included in the QCancer tool^9,10,12^, but it will also extend to additional symptoms from literature, PCUK website, and patient representatives. Data will be extracted for all relevant symptoms (e.g. dysphagia, abdominal pain, abdominal distension, appetite loss, weight loss, diarrhoea, constipation, tiredness, itching, back pain etc.) and investigations (e.g. anaemia, blood glucose, hormone levels) recorded in the database before the index date.

### 4.8 Potential risk factors

We will also identify potential risk factors based on existing research^9,10,12,15^, supplemented by an up-to-date literature review. This is likely to include age, ethnicity, socioeconomic deprivation, lifestyle factors (e.g. smoking, alcohol, body mass index), family history of cancer, previous diagnosis of other types of cancer, pancreatitis, diabetes, other comorbidities, prescribed medications, etc.

### 4.9 Statistical analysis

We will run a descriptive analysis first to compare the characteristics of patient groups

a. cases with PNENs;
b. cases with PDAC;
c. matched controls (without pancreatic cancer)

We will examine patients’ records to identify ‘red flag’ symptoms which occurred prior to the diagnosis of pancreatic cancer, or the equivalent date in matched controls. We will examine different time periods e.g. < 1 months; 1-3 months; 4-6 months; 7-12 months; 13-24 months and > 24 months before diagnosis. A univariate analysis will be conducted to examine each candidate symptom and risk factor in turn, selecting those significantly associated with PNENs and PDAC. We will then include all the red flag symptoms in a multivariate analysis using **conditional logistic regression**. This will determine which red flag symptoms remain important and significant when all the factors are taken into account simultaneously. This process along with statistical tests of interaction will help identify whether the red flag symptoms for PNEN are similar or different from the red flag symptoms for PDAC. It will also help identify relevant time periods prior to diagnosis.

#### Power analysis and consideration

a feasibility study identified over 30,000 patients with pancreatic cancer of which around 620 (2.1%) had PNENs in the last 20 years. With 620 cases and 10 matched controls per case we will be able to detect an odds ratio of 1.89 or more for a symptom recorded in 5% of controls, with 90% power and 1% significance, assuming a correlation coefficient for exposure between matched cases and controls of 0.1. For a 1% exposure, the odds ratio is 3.23 or more.

#### Missing data

Missing data in some important patient characteristics such as ethnicity, Townsend quintile, body mass index (BMI), smoking and drinking statuses will be checked. Multiple imputation with chained equations will be used to impute missing values for these variables, under the assumption of missing at random (MAR). All other covariates and the outcome variables were included in the imputation model. Ten imputations will be carried out. Rubin’s rules will be used to pool the parameter estimates across the imputed datasets_16_.

#### Sensitivity analysis

cases and controls who have less than three years of computerised data before the index date will be excluded in sensitivity analysis. The results between the full data analysis and sensitivity analysis will be compared.

## Data Availability

This study used anonymised NHS patient records from the EMIS system. Due to the sensitive nature of data, and to guarantee the confidentiality of personal and health information, only the authors have access to the data during the study in accordance with relevant license agreements.

## 6 Research Ethics Committee approval

This study utilising QResearch data has obtained approval from the QResearch Scientific Committee in July 2018 (Research ethics committee reference: 18/EM/0400). QResearch is a Research Ethics Approved Research Database, confirmed from the East Midlands – Derby Research Ethics Committee. Therefore, this also constitutes research ethics approval. For more information, please see: https://www.qresearch.org/about/ethics-and-confidentiality/#h-H3_2

## References

1. UK CR. Pancreatic cancer statistics - Key Facts2011. http://info.cancerresearchuk.org/cancerstats/keyfacts/pancreatic-cancer/ (accessed.

2. Li D, Xie K, Wolff R, Abbruzzese JL. Pancreatic cancer. Lancet 2004; 363(9414): 1049–57.

3. Bowles MJ, Benjamin IS. Cancer of the stomach and pancreas. BMJ 2001; 323(7326): 1413–6.

4. Purohit V, Khalsa J, Serrano J. Mechanisms of alcohol-associated cancers: introduction and summary of the symposium. Alcohol 2005; 35(3): 155–60.

5. Gullo L. Diabetes and the risk of pancreatic cancer. Ann Oncol 1999; 10 Suppl 4: 79–81.

6. Takhar AS, Palaniappan P, Dhingsa R, Lobo DN. Recent developments in diagnosis of pancreatic cancer. BMJ 2004; 329(7467): 668–73.

7. Richards MA. The National Awareness and Early Diagnosis Initiative in England: assembling the evidence. British journal of cancer 2009; 101 Suppl 2: S1–4.

8. Hippisley-Cox J. Validity and completeness of the NHS Number in primary and secondary care electronic data in England 1991-2013.2013. Hippisley-Cox J. Validity and completeness of the NHS number in primary and secondary care: electronic data in England 1991-2013 http://eprints.nottingham.ac.uk/3153/1/Validity%26CompletenessNHSNumber.pdf (accessed June 2013).

9. Hippisley-Cox J, Coupland C. Symptoms and risk factors to identify men with suspected cancer in primary care: derivation and validation of an algorithm. Br J Gen Pract 2013; 63(606): 1–10.

10. Hippisley-Cox J, Coupland C. Symptoms and risk factors to identify women with suspected cancer in primary care: derivation and validation of an algorithm. Br J Gen Pract 2013; 63(606): 11–21.

11. Hippisley-Cox J, Coupland C. Development and validation of QDiabetes-2018 risk prediction algorithm to estimate future risk of type 2 diabetes: cohort study. BMJ 2017; 359.

12. Hippisley-Cox J, Coupland C. Identifying patients with suspected pancreatic cancer in primary care: derivation and validation of an algorithm. British Journal of General Practice 2012; 62(594): e38–e45.

13. Hippisley-Cox J, Coupland C. Development and validation of risk prediction equations to estimate survival in patients with colorectal cancer: cohort study. BMJ 2017; 357.

14. Richardson DB. An incidence density sampling program for nested case-control analyses. Occupational and Environmental Medicine 2004; 61(12): e59–e.

15. Hippisley-Cox J, Coupland C. Development and validation of risk prediction algorithms to estimate future risk of common cancers in men and women: prospective cohort study. BMJ Open 2015; 5(3).

16. Rubin D. Multiple imputation for non-response in surveys. New York, NY: John Wiley; 1987.

